# CHANGES IN CORNEAL ABERRATIONS IN PATIENTS WITH CATARACT SURGERY WITH TORIC LENS IMPLANT: AN EXPLORATORY REVIEW

**DOI:** 10.1101/2023.08.16.23294174

**Authors:** Jorge Karim Assis, Luisa Fernanda Montaño, DAniela Tutal Quintero, Simon Giraldo

**Affiliations:** San Martin University Foundation, Cali-Colombia

**Keywords:** Corneal Diseases, Corneal Wavefront Aberration, Lens Implantation, Intraocular y Ophthalmology

## Abstract

**Introduction:** The cataract is one of the main causes of visual alterations, which is normally treated with surgery since it improves the visual acuity of the patients, the object of the present investigation is to synthesize evidence on how the surgical intervention affects or changes the corneal aberrations.

**Methodology:** conducted an exploratory review of the literature, observational studies on changes in corneal aberrations after cataract surgery with toric lens implant, using Pubmed, Google Scholar, Lilacs, Scopus, and Science Direct, searching and selecting articles from blindly and independently, following the PRISMA methodology.

**Results:** The body of evidence selected was 4 articles published in a 6-year window, between 2011 and 2016. The average age of the surgically operated population was 65.3 ± 7.7 years, while the proportion according to sex there was a similar and greater change in corneal aberrations due to coma and trefoil.

**Conclusions:** surgery with an incision equal to or less than 2.2 mm induces slight changes in the aberration of the total cornea, which increase before in eyes with a high pre-existence of corneal astigmatism and it is independent if the lens is toric or not.

## INTRODUCTION

Visual health is key in all aspects and activities of human daily life, the eyes are the basis of the sense of sight, being a very delicate and highly complex organ; where one of the main problems are cataracts, which consists of an opacity in the lens of the eye, which has a multifactorial etiology that produces a slow and progressive decrease in vision. This can appear at any age, although it is more common in the elderly and is the most common cause of reversible vision loss worldwide (1).

According to global figures from the World Health Organization, it is estimated that around 300 million people in the world suffer from some type of visual impairment or blindness. Being the cataract the main cause of these alterations, representing 51% of the total. In the American continent, 26.8% of the population has some type of visual impairment, presenting 3,500 people with legal blindness for every million inhabitants, being more prevalent in adults older than 50 years where about 82% of the population suffers some alteration, a figure that has been increasing due to the aging population that humanity is going through (2).

Cataract surgery is one of the most performed procedures worldwide. Along with advances in cataract surgical techniques, there have been improvements in intraocular lens replacement technology. Cataract surgery can be considered one of the most successful treatments in all of medicine and is, up to now, the only possible treatment to improve the vision of the cataract patient. Surgical treatment of cataract has evolved over time from Couching dislocation to modern phacoemulsification and lens replacement (1,3).

In cataract surgery, the toric intraocular lens is currently used, which is a type of lens that is used to eliminate astigmatism in eye surgery. It is characterized because it has different optical power and focal length in two mutually perpendicular orientations that allows the lens to create different refractive and focusing powers when the eye is in a horizontal and vertical direction (4).

Optical aberrations are imperfections in an optical system that produce defective images and prevent the reproduction of a clear and exact copy of the fixation object. When analyzing, for example, an eye with any refractive error, it is found that the outgoing light beams lose their parallelism and some of them lead or lag with respect to the reference plane. This is etymologically called corneal or optical aberration or wavefront deformity, as shown in Figure 1.

**Figure 1.**
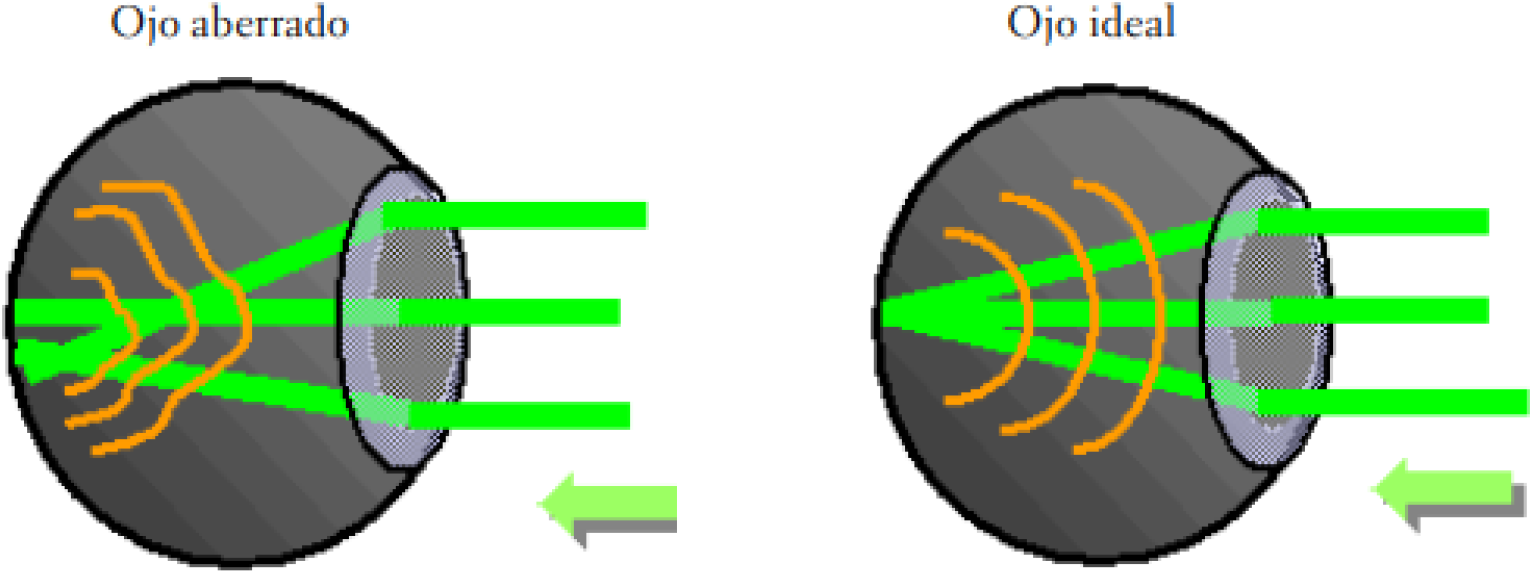
Perfect eye versus aberrated eye

Although cataract surgery improves the visual acuity of patients, a phenomenon that is little known and/or studied is how surgical intervention affects or changes corneal aberrations, for which the objective of the present investigation was to synthesize available evidence on the changes in corneal aberrations before and after intraocular cataract surgery with toric lens.

## METHODOLOGY

### Type of study

The present investigation carried out an exploratory review of the literature, with the objective of synthesizing currently available scientific evidence on the magnitude and distribution of corneal aberrations after cataract surgery with toric lens implant.

### Inclusion criteria

Articles published in indexed journals were included, which had the following characteristics:

- Performed on the adult population aged 18 and over
- Observational studies
- Of population with cataract surgery with toric lens implant
- To perform aberrometric measurements before and after cataract surgery by phacoemulsification with a toric lens implant.
- In English and Spanish

### Exclusion criteria

- Studies with diabetic and/or hypertensive patients, who present glaucoma, macular degeneration or who have endothelial counts less than 1200 cells.
- Studies with a low level of methodological quality according to the STROBE checklist.

### Information sources

The sources of information for the search for available evidence on the changes in optical aberrations in patients with cataract surgery with toric lens implant, were carried out using the main databases and search engines of scientific evidence at present (Table 1).

**Table 1.** Databases and search engines used.

| Database | URL | Start Date | End Date |
| --- | --- | --- | --- |
| Pubmed | https://pubmed.ncbi.nlm.nih.gov/ | 20/09/2021 | 30/09/2021 |
| Google Scholar | https://scholar.google.com/ | 30/09/2021 | 10/10/2021 |
| Lilacs | https://lilacs.bvsalud.org/es/ | 10/10/2021 | 20/10/2021 |
| Scopus | https://www.scopus.com/home.uri | 20/10/2021 | 30/10/2021 |
| Science direct | https://www.sciencedirect.com/ | 30/10/2021 | 10/11/2021 |

**Table 2.** PICO Structured Research Question Table.

| PICO Question |  |
| --- | --- |
| P – Population: | Over 18 years |
| I - Intervention: | Phacoemulsification cataract surgery + toric lens implant |
| C – Comparison: | Does not apply |
| O – Outcome: | Changes in corneal aberrations |

**Table 3.** Search strategy for database.

| Database | Search strategy | Filter |
| --- | --- | --- |
| Pubmed | 1. Toric intraocular lens AND Corneal Wavefront Aberration | Observational studies, humans, adult (18+ years) |
| Google scholar | 1. Corneal Wavefront Aberration "Toric intraocular lens" |  |
| Lillacs | 1. Implantación de Lentes Intraoculares [Palabras] and Aberración de Frente de Onda Corneal [Palabras] |  |
| Scopus | 1. ALL ( "Toric intraocular lens" ) AND ALL ( "Corneal Wavefront Aberration" ) |  |
| Science direct | 1. "Toric intraocular lens" AND Corneal Wavefront Aberration |  |

### Search Strategy

The search strategy was raised according to the clinical research question, through the peak strategy, as evidenced below:

### Selection process

The selection process was carried out blindly and independently by the two researchers, hereinafter EDT and LFM, using the typing criteria of numeral 5.2. At the end of the screening and selection process, the final articles were compared, adjusting the differences by part of an argumentative process; In case of not reaching an agreement between the two researchers, an arbitration process was carried out by a third SAG researcher.

### Duplicates

The five searches carried out totaled (n=877 articles) were exported to Microsoft Excel 2013® through a .cvs file, where duplicates were automatically eliminated first by the DOI with the eliminate duplicates function and second manually through the revision of the title of each article; leaving a total of 769 unique items.

### screening

With the 769 unique articles, a screening process was carried out, reading the title and the abstract, to determine if the article met the typification criteria mentioned above in numeral 5.2, in this EDT and LFM process, a total of 716 were excluded. and 721 articles respectively; leaving 51 and 48 pre-selected items for the selection process.

### Selection

The selection process consisted of reading the entire article to confirm those studies preselected in the screening stage. In this process, EDT and LFM excluded a total of 39 and 37 articles, respectively. With the articles selected after complete reading, a critical reading was carried out using the STROBE checklist for observational studies, in order to establish the methodological quality of the selected articles, a process where EDT and LFM excluded a total of 2 and 3 articles. respectively, finally selecting 12 and 8 final articles.

When comparing the articles at this point, it was found that they differed from each other and added a total of 13 unique articles, in addition there were differences in the screening stages, selection by complete reading and by checklist, of which a comparison process was carried out. of articles thus leaving 9 final studies to carry out the qualitative synthesis.

### Data collection and analysis plan

In the collection stage, 5 articles were excluded because they only recorded the measurement of corneal aberration after surgery and not in the preoperative stage, which did not allow determining the change in corneal aberration before and after surgery, thus leaving 4 final articles. to develop the qualitative synthesis.

For the collection of information from the 4 selected articles, the guide tables of the manual for the preparation of systematic reviews of the Cochrane collaboration were taken into consideration.

For the qualitative synthesis, a flowchart was made according to the PRISMA methodology, where the process of conformation of the body of evidence was evidenced, documenting the volume of studies in each of the stages of search, screening and selection of articles.

A descriptive analysis of the year of publication, the origin of the authors, the patients, the number of participants, the number of eyes, the type of design, the type of intraocular lens implant surgery, the aberration measurement time, was performed. post surgery, the prevalence of corneal aberrations, the types of aberration found.

### Ethical Considerations

In accordance with resolution 8430 of 1993, which establishes the scientific, technical, and administrative standards for health research, and based on article 11 of this document, where it regulates and classifies them according to risk, this study would fall within an investigation. without risk, since it uses documentary analysis methods in the different search engines such as Pubmed, Scopus, Science direct, Lilacs, Google Scholar, so no intentional modification or alteration of the biological, physiological, psychological or social of any individual.

## RESULTS

Among the 877 consolidated articles, 108 duplicates were identified among the five databases or search engines used, thus leaving 769 unique articles that were screened by reviewing the title and the abstract to determine if the study met the classification criteria. (inclusion and exclusion), excluding in this phase a total of 716 articles, because 311 corresponded to a single case report or series of cases with less than 10 operated eyes, 108 were clinical trials, 191 were review articles (narrative) focused in other aspects of visual quality and 106 because the population had basic pathologies such as glaucoma or wavefront aberrations did not mediate, if not visual acuity and other measures, thus leaving a total of 51 articles for the selection process that would later After reading the article completely and applying the STROBE checklist, 39 articles were excluded because they did not measure corneal aberrations in microns before and after surgery, because they included a population that did not match the typing criteria of the present investigation and due to methodological criteria. ; later in the information extraction process, 8 studies were excluded because they did not have complete information or it was measured on scales or times that were not comparable for the study, thus leaving 4 studies to carry out the qualitative synthesis of results (Figure 2).

**Figure 2.**
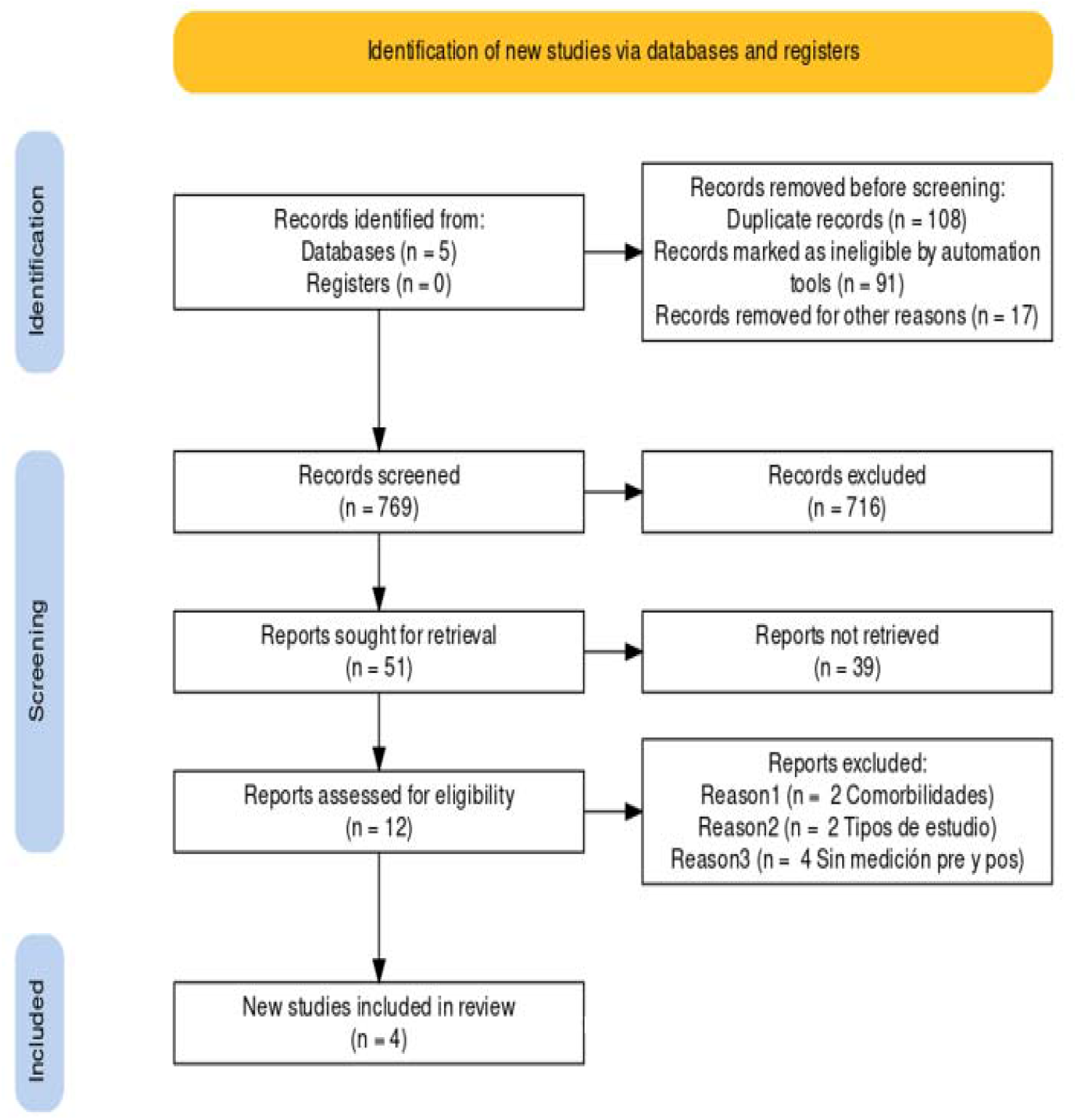
PRISMA flow chart.

The selected body of evidence was published in a 6-year window, between 2011 and 2016, the publication volume per year was homogeneous with a frequency of one annual publication, although 2013 and 2014 did not record any articles. The countries of the Asian and European continent such as Japan, Spain and the Czech Republic summarized the total number of publications, without finding any article carried out in Latin America.

Most of the investigations were carried out in Universities, Schools of Medicine or in Specialized Departments of Ophthalmology; The means of scientific dissemination is summarized in two journals, the Journal of Cataract & Refractive Surgery (JCRS) and The Japanese Journal of Ophthalmology, the first being a North American journal that has been published since 1986 and has an H index of 142, while the second is a Japanese magazine, which has been published since 1973 and has an H index of 56 (Table 4).

**Table 4.** Identification characteristics of the selected body of evidence.

| No | Authors | Title | Year of publication | Country | University/hospital | Journal |
| --- | --- | --- | --- | --- | --- | --- |
| 1 | Alió et al (27) | Vector analysis of astigmatic changes after cataract surgery with toric intraocular lens implantation | 2011 | Spain | Vissum-Instituto de Oftalmológico de Alicante, Alicante, Spain | Journal of Cataract & Refractive Surgery |
| 2 | Hayashi et al (28) | Higher-order aberrations and visual function in pseudophakic eyes with a toric intraocular lens | 2012 | Japan | Hayashi Eye Hospital, Fukuoka, Japón. | Journal of Cataract & Refractive Surgery |
| 3 | Mojzis et al (29) | Visual outcomes of a new toric trifocal diffractive intraocular lens | 2015 | Spain-Czech Republic | Regional Hospital in Havlickuv Brod - Universidad Miguel Hernandez | Journal of Cataract & Refractive Surgery |
| 4 | Hidaka et al (30) | Changes in corneal aberrations after cataract | 2016 | Japan | Keio University Hospital. | The Japanese |
|  |  | surgery |  |  |  | Journal of<br>Ophthalmol<br>ogy |

On average, 34 ± 11.7 people participated in each study with a range of 21 to 49, evidencing low variability in the population volume under study, an average of 35.5 ± 9.7 eyes were treated, with a range of 27 to 49 eyes. evidencing variability in the population volume under study, an average of 104 eyes were treated for every 100 participants; The average age of the surgically operated population was 65.3 ± 7.7 years, while the proportion according to sex was similar with an average of 50.7 ± 10% women, finding that of the four studies, two reported a greater volume. of the male population, in one female and in another the distribution by sex was not reported, all surgeries were performed by phacoemulsification, however the type of incision changed, finding that two performed microincision cataract surgery (MICS), one performed a superior sclerocorneal incision (SSI) and another clear corneal incision (CCI), the time of post-surgical measurement varied between 1 and 6 months (Table 5).

**Table 5.** Methodological characteristics of the selected body of evidence.

| No | Authors | Average age | % Women | patients | Eyes | Incision | Surgery Type | Time after surgery (months) |
| --- | --- | --- | --- | --- | --- | --- | --- | --- |
| 1 | Alió et al (27) | 70,05 | 61,9% | 21 | 27 | MICS | Phacoemulsification | 6 |
| 2 | Hayashi et al (28) | 70,3 | 42,9% | 49 | 49 | CCI<sup>1</sup> | Phacoemulsification | 6 |
| 3 | Mojzis et al (29) | 53,9 | - | 30 | 30 | MICS<sup>2</sup> | Phacoemulsification | 3 |
| 4 | Hidaka et al (30) | 67,3 | 47,2% | 36 | 36 | SSI<sup>3</sup> | Phacoemulsification | 1 |

<sup>1</sup> clear corneal incision, <sup>2</sup> microincision cataract surgery, <sup>3</sup> superior sclerocorneal incisión.

Among the four types of corneal aberrations evaluated, coma reported the greatest difference with a mean of 0.0582 ± 0.0432 microns, finding that the median dropped to 0.0400, indicating the existence of high changes in aberrations that raise the average. being caused by the differences found in the study by Mojzis et al (29), with a change before and after surgery of 0.1075 microns; The second place in magnitude of differences in the change of corneal aberrations was occupied by the trefoil with an average of 0.0445 ± 0.0286 microns, a median of 0.0300 again indicating the existence of high differences in aberrations that raise the average., being caused by the discrepancies found in the study by Mojzis et al (29), with a change before and after surgery of 0.0775 microns.

Regarding the variability in the change before and after surgery of the corneal aberrations, it was higher in the “coma” aberration followed by those of high order (HOA), in both said variabilities is due to the wide difference between the change registered in the study by Mojzis et al (29) which was 0.1075 and 0.0603 compared to the low difference in the study by Hidaka et al (30) 0.0270 and 0.0050 (Table 6).

**Table 6.** Absolute difference in microns of corneal aberrations before and after cataract surgery with toric lens implant in the selected body of evidence.

| No | Authors | spherical (μm) | Coma (μm) | HOA (μm) | Trefoil (μm) |
| --- | --- | --- | --- | --- | --- |
| 1 | Alió et al (27) | 0,0100 | 0,0400 | - | - |
| 2 | Hayashi et al (28) | - | - | 0,0550 | 0,0300 |
| 3 | Mojzis et al (29) | 0,0300 | 0,1075 | 0,0603 | 0,0775 |
| 4 | Hidaka et al (30) | 0,0120 | 0,0270 | 0,0050 | 0,0260 |

## DISCUSSION

The selected body of evidence was published in a 6-year window, between 2011 and 2016, the volume of publication per year was homogeneous and the countries of the Asian and European continent such as Japan, Spain and the Czech Republic summarized the total number of publications, without find no article made in Latin America, which reflects the high difference in publications from Latin America in the world’s indexed journals, which is not presented exclusively in ophthalmological issues but in everything that concerns health issues, contributing only approximately 3% of publications, since third world countries do not make a significant contribution to science, which is led by the great industrial powers, such as the US, Japan, the UK and Germany (31).

Most of the investigations were carried out in Universities, Schools of Medicine or in Specialized Departments of Ophthalmology; The means of scientific dissemination is summarized in two journals, the Journal of Cataract & Refractive Surgery (JCRS) and The Japanese Journal of Ophthalmology, the first being a North American journal that has been published since 1986 and has an H index of 142, while the second is a Japanese magazine, which has been published since 1973 and has an H-index of 56; showing a high difference

On average, 34 ± 11.7 people participated and 35.5 ± 9.7 eyes were treated. The average ratio was 104 eyes per 100 participants; The average age of the surgically operated population was 65.3 ± 7.7 years, while the proportion according to sex was similar with an average of 50.7 ± 10% women, finding that of the four studies, two reported a greater volume. of male population, in one female and in another the distribution by sex was not reported; all surgeries were performed by phacoemulsification, however the type of incision changed, finding that two performed microincision cataract surgery (MICS), one performed a superior sclerocorneal incision (SSI) and another clear corneal incision (CCI), the time of post-surgical measurement varied between 1 and 6 months.

Among the four types of corneal aberrations evaluated, “coma” reported the greatest difference with a mean of 0.0582 ± 0.0432 microns, finding that the median dropped to 0.0400, indicating the existence of high changes in the aberrations that raise the average, being caused by the differences found in the study by Mojzis et al (29), with a change before and after surgery of 0.1075 microns; The second place in magnitude of differences in the change of corneal aberrations was occupied by the trefoil with an average of 0.0445 ± 0.0286 microns, a median of 0.0300 again indicating the existence of high differences in aberrations that raise the average., being caused by the discrepancies found in the study by Mojzis et al (29), with a change before and after surgery of 0.0775 microns. Similar findings have been reported in various studies that have found that multifocal intraocular implant surgery induces considerable changes in high-order HOA aberrations, especially “coma”. (32–34), therefore, better IOL stability over time may cause less tilt or coma from small decentrations; however, these findings are not yet conclusive and should be addressed in future studies.

For their part, Hayashi et al (35) found that high-order and third-order aberrations such as the “trefoil” were significantly higher after surgery, mainly in eyes with a high pre-existence of corneal astigmatism; this change is predominantly attributed to to the difference in irregular corneal astigmatism induced or caused by the “clear corneal incision CCI” surgical incision. Being higher in the toric group and the high astigmatism group, although no significant differences were found using the multivariate analysis; an important finding of Hayashi is that the changes in aberrations are not due to the type of lens whether it is toric or non-toric, however, it is presumed that they can be induced by a marked misalignment of the axis of the toric IOL, although this statement requires further investigation.

In the selected body of evidence Mojzis et al (29) reported the greatest change in third-order corneal aberrations such as “coma” and “trefoil”; Regarding the analysis of the aberrometric results, this study only registered significant post-surgical changes in the Z_4^4 tetrafoil aberration, although this was not evaluated in the present investigation, in the rest of the aberrations no significant changes were evidenced before and after the surgery, confirming the stability of the corneal optics after surgery using a microincision technique. Detecting non-significant changes in internal or ocular aberrations. These findings demonstrate the low incidence of aberrations with the implantation of this toric trifocal IOL model after cataract surgery.

The mean change in high order aberrations (HOA) comparing before versus after cataract surgery with toric lens implant was 0.0401 ± 0.0305 μm microns, some studies have also reported similar changes in HOAs. (27,36-38). Regarding the post-surgical changes induced in the global change of the HOA, declaring a small incision, Tong et al. (38), although the change was not statistically significant (P = 0.099) for a 6-mm pupil diameter with a 3.0-mm incision. Elkady et al. (37) for their part, reported a change of 0.47 ± 0.26 μm before the operation and 0.59 ± 0.32 μm 1 month after the operation, reflecting a difference of 0.12 μm, without finding significant differences. (P=0.47) for a 6-mm pupil diameter with a 1.5-mm incision. On the other hand, Can et al. (39) reported that the higher order change was from 0.57 ± 0.24 μm to 0.62 ± 0.26 μm, resulting in a difference of 0.05 μm for a 6 mm diameter pupil across a 1.7 mm incision (P = 0.658). Alió et al. (37) found that the higher order change increased significantly (P = 0.01) from 0.45 ± 0.19 μm to 0.53 ± 0.29 μm, resulting in a difference of 0.08 μm, for a diameter 6 mm pupillary and a 2.2 mm incision. Like Guirao et al. (40) reported that the total change increased significantly (P = 0.05) from 0.66 ± 0.42 μm to 0.99 ± 0.80 μm, reflecting a difference of 0.34 μm for a 6-mm aperture diameter across through a 3.5 mm incision. It should be noted that none evaluated the corneal aberrations of the anterior corneal surface, these findings show that the changes in posterior corneal aberrations after cataract surgery with an incision of 2.2 mm or less are numerically irrelevant, in addition that it is considered that these changes in corneal aberrations are mainly due to the size and position of the incision, although there is not enough evidence to prove this.

Regarding the variability in the change before and after surgery of the corneal aberrations, it was higher in the coma aberration followed by the high order aberrations (HOA), in both said variability is due to the wide difference between the change registered in the study of Mojzis et al (29) which was 0.1075 and 0.0603 compared to the low difference in the study by Hidaka et al (30) 0.0270 and 0.0050.

### Conclusions

The mean change of the four types of corneal aberrations evaluated was higher in the years 2015-2016, in younger patients (53 years), with a lower percentage of women between 42 and 47%, with microincision cataract surgery and clear corneal incision and with aberration measurement 3 months after surgery.

In summary, the results of the body of evidence showed that surgery with an incision equal to or less than 2.2 mm induces slight changes in the aberration of the total cornea, which increase before in eyes with a high pre-existence of corneal astigmatism., this change is predominantly attributed to the difference in irregular corneal astigmatism induced or caused by the surgical incision and is independent of whether the lens is toric or not.

### limitations

One of the main limitations in the present investigation lies in the weakness in the thematic management of corneal aberrations, however, it had an external ophthalmologist who supported the thematic development of the present investigation; Another limitation lies in the absence of a librarian or similar expert in bibliographic searches; however, there was an epidemiologist with experience in database searches and reference management who advised on the development of the search and the collaboration guidelines were followed. Cochrane.

Regarding the body of evidence selected, the main limitations were that changes in corneal aberrations were not measured according to the effect of corneal diameter, corneal thickness, axial length, IOL implant, incision tunnel length, and eccentricity in each eye. although it is considered that the influence of these factors is probably low.

One of the limitations is that not all the individual Zernicke terms could be evaluated due to the inadequate repeatability of the aberrometry measurements using the Pentacam, which justifies future studies using other devices.

Another limitation corresponds to not evaluating the effect of the toric IOL component on ocular HOAs, particularly when there is marked misalignment of the toric IOL axis.

## Data Availability

All data produced in the present work are contained in the manuscript

